## Supplemental Figures for "Human Microbiome Mixture Analysis using Weighted Quantile Sum Regression"

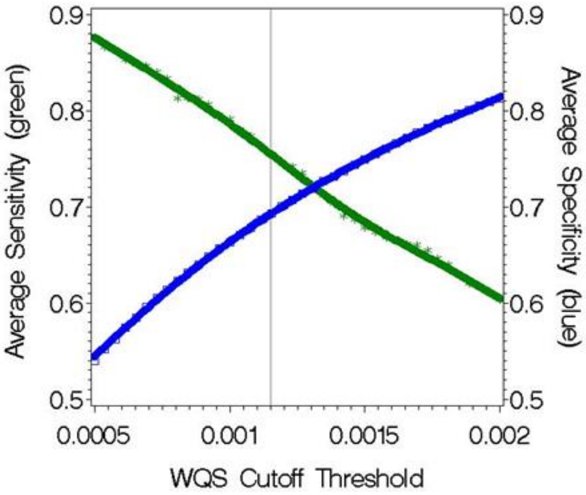


**Supplemental Fig 1**: **WQS Sensitivity and Specificity.** Average sensitivity and specificity based on a range of threshold cutoff values from 30 repeated holdout analyses for the Weighted Quantile Sum (WQS) operational taxonomic unit (OTU) index including 868 components.

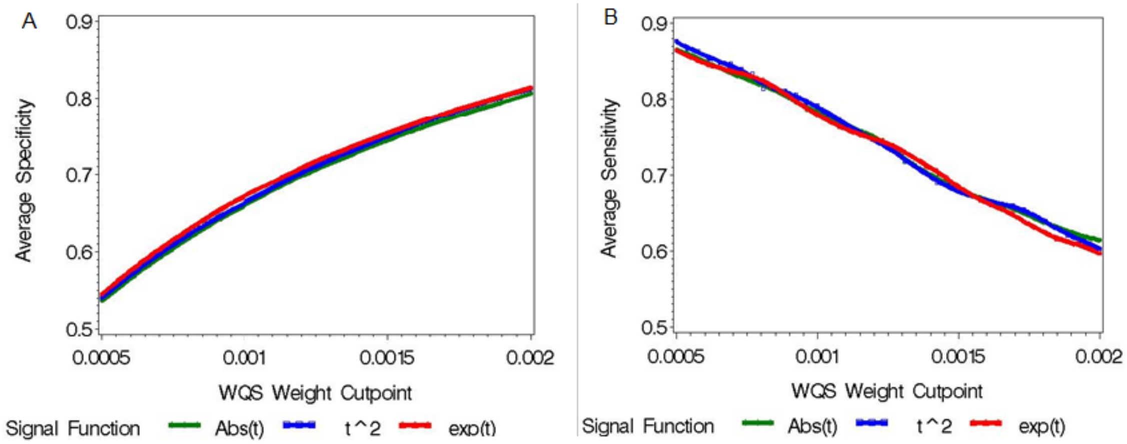


**Supplemental Fig 2**: LOESS plots of the average (A) specificity; and (B) sensitivity from the 30 repeated holdout datasets, across cutpoints and signal functions.
